# COVID-19 and Socioeconomic Factors: Cross-country Evidence

**DOI:** 10.1101/2020.10.22.20217430

**Authors:** Vishalkumar Jani, Dileep Mavalankar

**Affiliations:** Indian Institute of Public Health Gandhinagar (INDIA)

## Abstract

**Background:** COVID-19 pandemic has affected all countries across the globe in varying intensity resulting in varied numbers for total cases and deaths.

**Objectives:** The paper aims to understand if different socioeconomic factors have a role to play in determining the intensity of COVID-19 impact.

**Methods:** The study uses a country-wise number of corona cases and deaths and analyse them in a cross-country multivariate regression framework. It uses gross domestic product per capita, average temperature, population density, and median age as independent variables. The study uses testing data as a control variable.

**Results:** In absence of the testing variable, higher-income countries have experienced a higher number of COVID cases. The population density, median age, climate do not have significant impact. The countries with higher population density have lower deaths. Each region shows different patterns of correlation between socioeconomic factors and COVID intensity.

**Conclusion:** The majority of the cross-country variation can be attributed to the number of tests done by a country. The countries with high population density would have applied strict lockdowns and proactive testing to curb the deaths. The study essentially refutes claims around corona being a high-income group disease, cold-climate disease, or a disease impacting old-age patients more.

---

COVID-19 started in China towards the end of 2019 and has affected all countries across the globe in various intensity. World Health Organization (WHO) declared it to be a global pandemic in March, 2020.^1^ Majority of countries in Europe, Asia, and the Americas have witnessed an increasing number of total cases and deaths due to COVID in the last few months. However, it is important to analyse the country-level differences in the COVID experience in terms of total cases and deaths in the context of socioeconomic conditions.

Each country has tried to manage the pandemic with differing approaches and measures based on the resources and strength of the health system each has. However, there are certain underlying assumptions in adopting these different measures. However, due to various economic and social pressures, different country could implement containment measures with differing intensity.^2,3,4^

There are certain assertions made early in the pandemic about the age profile of the country, population density, climatic condition, or the development status of the country to moderate the impact caused by the COVID pandemic. The initial level data analysis about these factors and it was found that advanced age and higher unemployment rates increase the total cases of COVID.^2^

The main aim of the paper is to understand what country-specific socioeconomic variables do impact the number of cases and deaths in a country due to COVID. This paper is to advance a similar analysis using other important variables like population density, income level of the country, the number of tests done in a country, and the average temperature a country experience. The study also analyzes the cross-country data for the regional (continent wise) and human development groups wise analysis.

## Data and Methodology

The study uses 167 countries data as of October 15, 2020, from publicly available sources like the WHO database,^5^ Centre for Disease Control and Prevention database,^6^ and Worldometer COVID data site.^7^ The other socioeconomic indicators like gross domestic product (GDP) per capita, and population density have been sourced from the World Bank Development Indicators database.^8^ The Human Development Report 2019^9^is used for human development status, and the Weather-Atlas web database^10^ is used for average temperature data across countries.

The study uses a multivariate ordinary least squares regression method for analyzing the cross-sectional country-level data. It is acknowledged that this method may not be sufficient to establish causality, but it is efficient in identifying relationships between variables. The model is in a log-log format to estimate the elasticity between the variables under study. The study uses robust standard errors to make inferences about the significance of the regression coefficients.

The analysis includes two levels of analysis based on the dependent variable form: total numbers and per million population numbers. As and when, total cases or total deaths are used as dependent variables, COVID testing data of the total number of tests conducted is used, and when the dependent variable is in per million form, tests per million population is used as independent variable.

## Results

The main aim of the study is to decipher the relation between country-specific socio-economic indicators on the COVID cases and deaths.

Table 1 shows the result about total cases and deaths across all countries. The first two columns show the impact of various socioeconomic indicators on the total cases in a country, and the last two columns do the same for total deaths happened in a country due to COVID. Model 1 indicates that the countries with higher GDP per capita have experienced a higher number of COVID cases. However, model 2 shows that in presence of total number of tests done as an independent variable, this relationship between GDP per capita and number of cases vanishes. The adjusted R-squared of Model 2 indicates that it explains significantly higher variation in the total cases compared to that of Model 1. It is apparent that the number of tests done has a significant impact on the total number of cases at the country level.

**Table 1:** Total cases and Deaths and socio-economic determinants.

|  | (1)<br>ln (Total<br>cases) | (2)<br>ln (Total<br>cases) | (3)<br>ln (Total<br>deaths) | (4)<br>ln (Total<br>deaths) |
| --- | --- | --- | --- | --- |
| ln (GDP per capita) | 0.532**<br>(0.259) | 0.0031<br>(0.210) | 0.484<br>(0.266) | -0.296<br>(0.246) |
| ln (Population density) | 0.112<br>(0.128) | 0.143<br>(0.119) | -0.025<br>(0.154) | -0.373**<br>(0.152) |
| ln (Median age) | 0.054<br>(1.216) | 1.537*<br>(0.827) | 0.298<br>(1.197) | 0.324<br>(1.011) |
| ln (Average temperature) | 0.134<br>(0.432) | 0.264<br>(0.322) | 0.498<br>(0.468) | 0.085<br>(0.419) |
| ln (Total tests) |  | 0.987***<br>(0.0957) |  | 1.004***<br>(0.0977) |
| constant | 4.319<br>(3.318) | 2.493<br>(2.394) | 2.876<br>(3.412) | -1.723<br>(3.224) |
| N | 167 | 118 | 159 | 116 |
| Adjusted R-square | 0.079 | 0.623 | 0.072 | 0.506 |

Note: Standard errors in the parentheses; \* p< 0.1, \*\* p<0.05, \*\*\* p<0.01

Model 3 and Model 4 from Table 1 also show similar results that most of the variation in the total deaths across the country is explained by the number of tests a country has conducted. The population density does not have any significant impact on the number of cases detected but it has a statistically significant impact on the total deaths in a country. This means that as the population density of a country increases by one percentage, the deaths due to COVID are reducing by 0.37%.

Table 2 shows the same analysis as Table 1 with the dependent variables in per million form. This analysis takes care of the scaling to the population size. It shows the similar results as Table 1: most of the variation in COVID cases per million population or deaths per million population is significantly correlated with the tests per million population. The result of higher population density resulting in the lower deaths per million also holds statistically.

**Table 2:** Cases and Deaths per million and socioeconomic determinants.

|  | (1)<br>ln (Cases per<br>million) | (2)<br>ln (Cases per<br>million) | (3)<br>ln (Deaths per<br>million) | (4)<br>ln (Deaths per<br>million) |
| --- | --- | --- | --- | --- |
| ln (GDP per capita) | 0.764***<br>(0.202) | 0.256<br>(0.213) | 0.553***<br>(0.195) | 0.218<br>(0.247) |
| ln (Population density) | -0.025<br>(0.0894) | -0.143<br>(0.114) | -0.178*<br>(0.101) | -0.292**<br>(0.141) |
| ln (Median age) | 0.422<br>(0.961) | 1.266<br>(0.856) | 0.857<br>(0.947) | 0.071<br>(1.039) |
| ln (Average<br>temperature) | 0.127<br>(0.307) | 0.139<br>(0.316) | 0.184<br>(0.329) | 0.154<br>(0.390) |
| ln (Total tests) |  | 0.695***<br>(0.113) |  | 0.483***<br>(0.142) |
| constant | 1.415<br>(2.418) | 2.285<br>(2.268) | 3.199<br>(2.590) | 1.459<br>(2.865) |
| N | 167 | 118 | 158 | 115 |
| Adjusted R-square | 0.27 | 0.42 | 0.26 | 0.31 |

Note: Standard errors in the parentheses; \* p< 0.1, \*\* p<0.05, \*\*\* p<0.01

Tables 3 and 4 show the regression results that are specific to a particular region. Table 3 is about total COVID cases, and Table 4 uses total deaths as the dependent variable. This analysis is done to capture the regional differences that are masked by the regression models using a global sample.

**Table 3:** Region-wise total cases and socioeconomic determinants.

|  | Asia | Africa | Europe | North<br>America | South<br>America |
| --- | --- | --- | --- | --- | --- |
|  | ln (Total<br>cases) | ln (Total<br>cases) | ln (Total<br>cases) | ln (Total<br>cases) | ln (Total<br>cases) |
| ln (GDP per capita) | 0.678**<br>(0.319) | 0.583<br>(0.420) | -1.243***<br>(0.245) | -0.069<br>(1.283) | -3.068<br>(1.845) |
| ln (Population density) | 0.216<br>(0.186) | -0.694***<br>(0.241) | 0.433*<br>(0.247) | -0.794<br>(0.688) | 0.364<br>(0.601) |
| ln (Median age) | -5.813***<br>(1.583) | 3.526<br>(2.043) | 1.968<br>(2.827) | 3.548<br>(3.244) | -1.805<br>(5.115) |
| ln (Average temperature) | -0.634<br>(0.602) | 0.168<br>(1.516) | -0.867<br>(0.671) | 0.275<br>(0.262) | -0.496<br>(0.941) |
| ln (Total tests) | 0.805***<br>(0.231) | 0.952***<br>(0.0857) | 1.104***<br>(0.138) | 1.934***<br>(0.229) | 1.683***<br>(0.176) |
| constant | 13.59***<br>(4.527) | 6.575<br>(7.806) | 15.43*<br>(8.294) | -43.72<br>(20.19) | 31.75**<br>(7.624) |
| N | 33 | 25 | 39 | 9 | 10 |
| Adjusted R-square | 0.493 | 0.723 | 0.697 | 0.948 | 0.829 |

Note: Standard errors in the parentheses; \* p< 0.1, \*\* p<0.05, \*\*\* p<0.01

**Table 4:** Region-wise total deaths and socioeconomic determinants.

|  | Asia | Africa | Europe | North America | South America |
| --- | --- | --- | --- | --- | --- |
|  | ln (Total deaths) | ln (Total deaths) | ln (Total deaths) | ln (Total deaths) | ln (Total deaths) |
| ln (GDP per capita) | 0.068<br>(0.343) | 1.034<br>(0.628) | -1.472***<br>(0.421) | 1.135<br>(1.104) | -6.258*<br>(2.427) |
| ln (Population density) | -0.224<br>(0.191) | -0.713*<br>(0.351) | 0.642<br>(0.438) | 0.247<br>(0.602) | 1.010<br>(0.893) |
| ln (Median age) | -4.624**<br>(1.742) | -4.389*<br>(2.471) | -0.265<br>(3.848) | 1.445<br>(5.230) | 4.456<br>(6.057) |
| ln (Average temperature) | -0.734<br>(0.643) | -0.884<br>(2.160) | -1.167<br>(0.914) | 1.524<br>(1.234) | -1.641<br>(1.475) |
| ln (Total tests) | 0.935***<br>(0.207) | 0.756***<br>(0.115) | 1.234***<br>(0.213) | 2.675***<br>(0.351) | 1.628***<br>(0.170) |
| Constant | 12.00**<br>(4.628) | 6.989<br>(9.586) | 3.979<br>(11.70) | 75.12<br>(51.88) | 41.03**<br>(9.562) |
| N | 32 | 25 | 39 | 8 | 10 |
| Adjusted R-square | 0.524 | 0.481 | 0.599 | 0.864 | 0.763 |

Note: Standard errors in the parentheses; \* p< 0.1, \*\* p<0.05, \*\*\* p<0.01

Table 3 shows that, for all regions, the number of tests conducted is the most significant variable determining the total number of cases. Asian countries experience higher cases if the GDP per capita is high for a country. The model also shows that if the median age of the population increases by one percentage, the total cases may reduce by 5.7%. This is counterintuitive with the common understanding of higher age persons being more susceptible to the COVID. In the case of Europe, the per capita income has a negative relationship with the number of COVID case in a country. Moreover, the European country with higher population density have also reported a higher number of cases. This conflicts with the result shown by the African region, where population density and total number of cases are negatively correlated. In North or South America, no variable except the number of tests have a statistically significant relationship with the number of cases. Moreover, the number of North- and South American countries in the sample is too low, and hence it is best to leave them out of any inference drawing exercise.

Table 4 shows the result for total deaths in a country divided by the regions. The common factor impacting the number of deaths across different countries is the number of tests done. Apart from that Asia and Africa show that countries with higher median age has witnessed lowed total deaths. In addition to this, African countries also show that there is a negative relationship between population density and COVID deaths in a country. European countries show that there is a negative relationship between GDP per capita and the number of deaths due to COVID. This is in coherence with the lower number of cases with higher GDP per capita shown by Europe in Table 3.

## Discussion

The analysis of cross-country data available for COVID pandemic cases and deaths showed a peculiar pattern that most of the variation across the countries is explained by the number of tests conducted in a country. All other variables like per capita income or median age do not show any statistically significant impact on the total number or per million numbers of cases and deaths due to COVID.

The above overarching finding does suggest to the very important role of proactive testing using a thorough screening measure and policy. Many of the countries that did not have a very high number of cases initially have started witnessing increasing cases and deaths due to ramped-up testing. India is one prominent example of this. So, till the time a cure or vaccine is not found, it is essential to screen, trace, and identify the cases.

One of the most interesting findings of the analysis is countries with highly dense population seems to have lower deaths. This is counter-intuitive to the basic understanding of more density resulting in more spread of infection and a higher number of deaths due to the higher number of cases. However, this is not happening. Possibly the countries like India that have high population density ended up implementing a very strong lockdown in the initial phase of the pandemic. This resulted in heightened awareness and a more active community response to the situation and the chain of infection spread was broken. Moreover, the highly-dense populated areas in urban areas and they may have better health facilities to approach and active management by the local government may have averted a huge number of deaths.

A similar argument could be made about countries with higher median age in Asia and Africa having a lower number of cases and deaths due to COVID. As both of these regions are mainly developing countries, so low median age may be a result of the poor health system and hence may have resulted in more infections and more deaths. In the same way, the countries with the highest median age like Japan and South Korea either totally avoided the pandemic or controlled the spread through active screening and identification of cases very early.

Though the study is based on a very rudimentary data and methodology, it has important takeaways. In nutshell, the impact of the virus is homogenous across countries and regions and most of the variation across countries is explained by the testing numbers. It is a clear indication for active screening and testing. To flatten the curve, the screening curve has to increase at a higher rate compared to that of the spread of infection.

## Data Availability

We do not wish to make data public as all the data used is readily available in public domain.

## Notes

### Competing Interest Statement

The authors have declared no competing interest.

### Clinical Trial

Not applicable as this is a secondary data based research and not a clinical trial

### Funding Statement

No funding was required.

### Author Declarations

Exempted as the study is based on secondary publicly published data.

